# Early Detection of COVID-19 Outbreaks Using Human Mobility Data

**DOI:** 10.1101/2021.05.20.21257557

**Authors:** Grace Guan, Yotam Dery, Matan Yechezkel, Irad Ben-Gal, Dan Yamin, Margaret L. Brandeau

## Abstract

**Background:** Contact mixing plays a key role in the spread of COVID-19. Thus, mobility restrictions of varying degrees up to and including nationwide lockdowns have been implemented in over 200 countries. To appropriately target the timing, location, and severity of measures intended to encourage social distancing at a country level, it is essential to predict when and where outbreaks will occur, and how widespread they will be.

**Methods:** We analyze aggregated, anonymized health data and cell phone mobility data from Israel. We develop predictive models for daily new cases and the test positivity rate over the next 7 days for different geographic regions in Israel. We evaluate model goodness of fit using root mean squared error (RMSE). We use these predictions in a five-tier categorization scheme to predict the severity of COVID-19 in each region over the next week. We measure magnitude accuracy (MA), the extent to which the correct severity tier is predicted.

**Results:** Models using mobility data outperformed models that did not use mobility data, reducing RMSE by 17.3% when predicting new cases and by 10.2% when predicting the test positivity rate. The best set of predictors for new cases consisted of 1-day lag of past 7-day average new cases, along with a measure of internal movement within a region. The best set of predictors for the test positivity rate consisted of 3-days lag of past 7-day average test positivity rate, along with the same measure of internal movement. Using these predictors, RMSE was 4.812 cases per 100,000 people when predicting new cases and 0.79% when predicting the test positivity rate. MA in predicting new cases was 0.775, and accuracy of prediction to within one tier was 1.0. MA in predicting the test positivity rate was 0.820, and accuracy to within one tier was 0.998.

**Conclusions:** Using anonymized, macro-level data human mobility data along with health data aids predictions of when and where COVID-19 outbreaks are likely to occur. Our method provides a useful tool for government decision makers, particularly in the post-vaccination era, when focused interventions are needed to contain COVID-19 outbreaks while mitigating the collateral damage of more global restrictions.

## 1 Introduction

The global spread of SARS-CoV-2, the virus that causes COVID-19, has brought about the worst public health crisis in a generation. As of April 2021, there have been nearly 150 million COVID-19 cases and more than 3.1 million COVID-19 deaths (1, 2). To curb the spread of the virus, many countries have implemented interventions such as social distancing requirements, school closures, prohibitions on public gatherings, and complete lockdowns, with varying degrees of success (3–6). While lockdowns stymie COVID-19 transmission, drastic measures to encourage physical distancing may also cause social and economic harm (6, 7). Moreover, some interventions have come too little or too late, causing unnecessary hospital admissions and deaths (8, 9). Thus, early detection and prompt action are needed to contain outbreaks and minimize economic damage. It is also important to determine the appropriate severity of mobility restrictions: a full lockdown may not be called for when there is a minor outbreak, whereas minor social distancing restrictions will be insufficient if there is a major outbreak.

To appropriately target the timing, location, and severity of such measures, it is essential to be able to predict when and where outbreaks will occur and how widespread they will be. Recent work has focused on using past health data to forecast accumulated COVID-19 cases and deaths using exponential smoothing models (10, 11), autoregressive moving average models (11–13), and deep learning models (13–17).

One potentially useful input to such forecasts is human mobility data in the form of cell phone data (18, 19). Past studies have successfully used mobile phone geolocation data to predict the spatial spread of cholera and malaria (20, 21). Several studies have found relationships between mobile phone data and the spread of COVID-19 (22–24). However, these studies have not used the mobile phone data to make predictions about the trajectory of the COVID-19 epidemic.

Several studies have incorporated aggregated and anonymized mobile phone geolocation data into epidemic models to analyze the spread of COVID-19. One study used mobile phone data indicating “time spent at home” in an SEIR model (25), demonstrating that pandemic-induced decreases in mobility, as reflected by time spent at home, significantly reduced transmission of the virus in the U.S. Another study combined an SEIR model with a mobility network generated from mobile phone data (26). The analysis identified certain superspreader points inside cities that led to COVID-19 outbreaks, and then estimated the potential impact of closing various gathering places (e.g., restaurants, fitness centers, grocery stores). A study in Brazil used mobile phone data to predict the spatial-temporal spread of COVID-19 infections in cities within the states of São Paolo and Rio de Janeiro (27). The authors modeled the spread of infection within each city with an SI model, and captured travel between cities using cell phone data. Due to constant updates in measures and changes in individuals’ behavior (e.g., face masks, physical distancing regulations, shelter-in-place restrictions), the use of an epidemic models for prediction necessitates calibration to realized data at each stage of the epidemic in each region.

We take an alternate approach to prediction: we do not reflect transmission explicitly but instead use a model that learns from past health and mobility data. Our work adds to the literature on how human mobility drives disease transmission and is the first to use mobility data directly to predict daily new cases and the COVID-19 test positivity rate in each region of a country over time. We find that aggregated, anonymized human mobility data improves predictions of new COVID-19 cases and the test positivity rate, and classifications of outbreak severity, beyond predictions made using only past health data.

## 2 Methods

Using aggregated, anonymized health data and cell phone mobility data, we forecast next 7-day averages of new cases of COVID-19 and the COVID-19 test positivity rate for different geographic regions of Israel from March to December 2020. We combine these forecasts with a categorization system to predict the severity of COVID-19 in each region one week in advance.

Our work is motivated by the link between mobility and epidemic severity. Figure 1 highlights several regions of Israel over consecutive two-week periods in late 2020, with arrows denoting significant travel between regions in each period and shading indicating the transmission level. Regions that experienced inflows of people from high-transmission districts had higher transmission levels in future periods. For example, in the period beginning on November 12, the transmission level in the Golan Heights (upper right district in panel A) was high, and there was significant travel from this district to the Akko and Yizre’el districts (upper two districts marked by arrows in panel B). In the next time period, Akko and Yizre’el saw an increase in cases.

**Figure 1:**
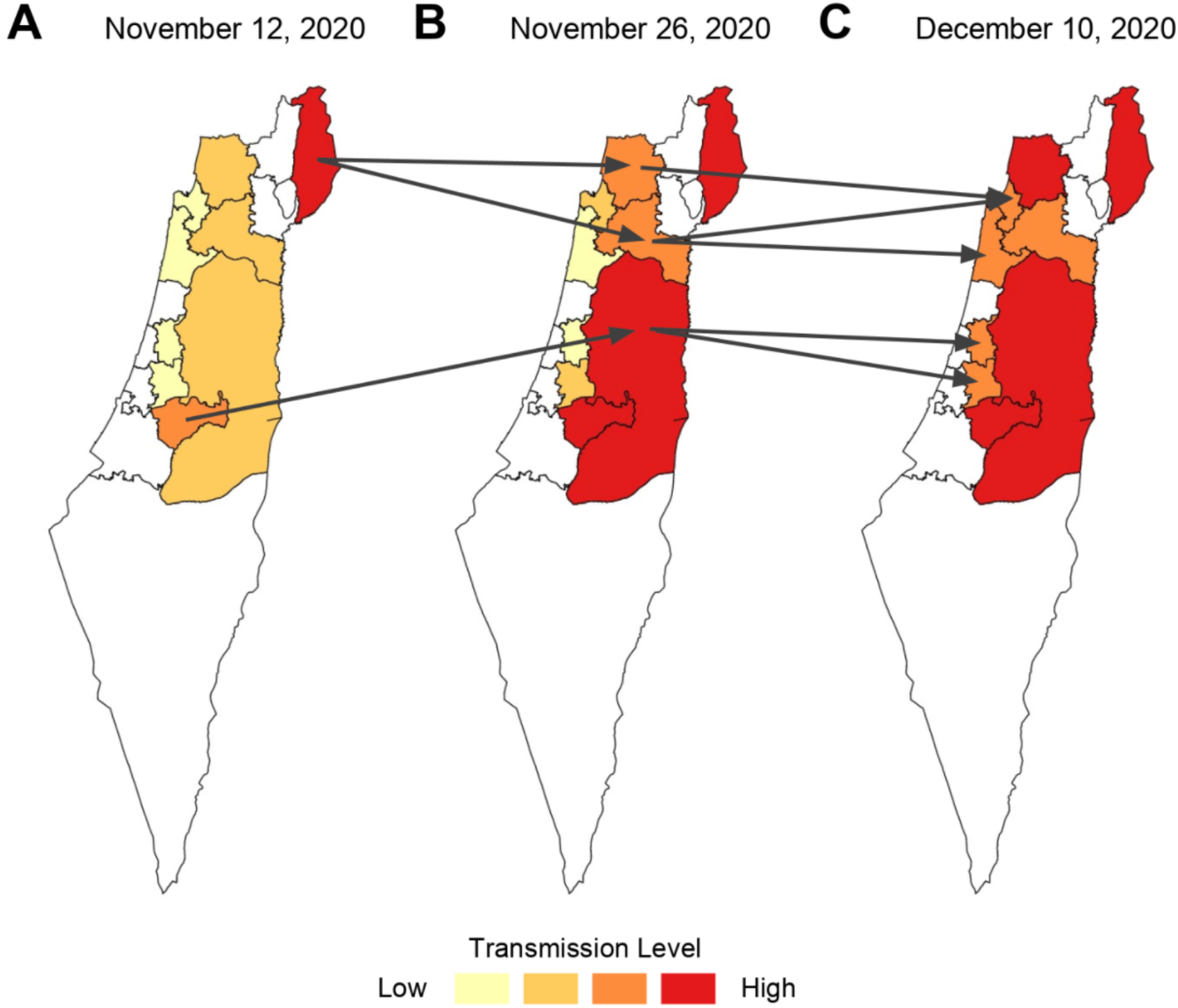
Association between movement between districts (arrows) and epidemic severity (indicated by shading) for two twerweek periods in late 2020. Arrows denote significant travel between regions over the two-week period.

### 2.1 Data

#### 2.1.1 Health data

We used publicly available, daily data on accumulated COVID-19 cases, active cases, tests, and recoveries from the Israeli Ministry of Health from February 1, 2020 to January 7, 2021 (28). Individuals were considered to have COVID-19 if they tested positive in an RT-PCR test. Individuals were deemed to be recovered either ten days after a positive test or three days after COVID-19-related symptoms disappear (excluding loss of smell and persistent cough), whichever is greater (29). Concurrent work has estimated that cases in this dataset are active on average for 8 days (30, 31). The dataset is stratified by 1,642 statistical regions comprising Israel as defined by the Israeli Central Bureau of Statistics.

We mapped the 1,642 statistical regions to 16 districts (Israeli “nafot”, Figure S.1). Our use of these larger districts neutralizes the noise in the lower-level data and allows for more accurate predictions.

We computed daily prevalence, new cases, new tests, and test positivity rate from this data. We normalized new cases and new tests by the population size of each district to be in terms of 100,000 people; “new cases” and “new tests” in the remainder of the paper refer to these population-adjusted values.

For privacy purposes, districts were included in our dataset when at least one statistical region in the district had accumulated at least 15 cases, tests, and recoveries. Seven-day average new cases for each district are displayed in Figure S.1. Most districts had similar trends. In our analysis we excluded one district (district 71) because the health data followed a pattern not seen by any other district (Figure S.1, pink line dominating all others). We excluded another district (district 29) that had fewer than 50,000 residents, far smaller than the average district size, as the data were not sufficient for prediction. The two excluded districts collectively contained 5.3% of Israel’s population; thus, our analysis focuses on 14 of the 16 districts, covering 94.7% of Israel’s population.

#### 2.1.2 Mobility Data

We used aggregated, anonymized cell phone mobility data from February 1, 2020 to November 30, 2020. This dataset was obtained from an Israeli cell phone carrier and is based on over 3 million Israeli cell phone users who are demographically, ethnically, and socioeconomically representative of the Israeli population. The dataset comprises daily and hourly movement patterns within and between 2,630 traffic analysis zones (TAZ regions, defined by the Israeli Ministry of Transportation) covering all of Israel. The movement patterns take the form of origin-destination (OD) data representing daily and hourly numbers of trips between pairs of regions. For privacy considerations, if fewer than 50 individuals were moving from one region to another in a given hour, the number of reported individuals was set to 1. We replaced these 1’s with a more realistic value of 7 in our analysis (32). Because the mobility dataset ended on November 30, 2020, for the next 31 days from December 1, 2020 to December 31, 2020, we used perturbed estimates based on mobility data from earlier dates during the pandemic that had similar restrictions in effect (details in Supplement A).

We mapped the 2,630 TAZ regions to the 16 districts for analysis.

#### 2.1.3 Socioeconomic and demographic data

For each district, we obtained 2017 data on the age distribution, the population size, and socioeconomic score from the Israeli Central Bureau of Statistics (CBS). The socioeconomic score is an integer value between 1 and 10, with 1 reflecting the most impoverished regions and 10 reflecting the wealthiest regions. The socioeconomic score is calculated by the CBS from demography, education, employment, and standard of living. Lastly, we calculated the median age for each district from the age distribution.

### 2.2 Model Inputs and Outputs

#### 2.2.1 Model Inputs: Developing predictors

We use 7-day average lagged health features to reflect COVID-19 incidence in each district over the past week. In addition to these solely health-based predictors, we combine health and mobility data to develop predictors reflecting the spread of COVID-19 between and within districts.

##### Lag features

Due to the availability of daily updated data, when predicting values for the next week, our models can see the previous days’ realized values for incidence and the test positivity rate. Thus, we introduce lag features, which allow pure time series problems to be converted into supervised learning problems. For example, a 1-day lag would have the label of day *X* – 1 input as a predictor for predicting on day *X*. Similarly, a 6-day lag would have the labels of days [*X* – 6, *X* – 5,…, *X* – 1] as six different predictors for predicting on day *X*. We test 1-day, 3-day, and 6-day lagged features for each prediction task.

##### Pressure Score

The Pressure Score for a district reflects potential cases imported into that district over the past *n* days. This metric has been used in the literature to predict the spatial spread of cholera with mobile phone data (20). For each day *t*, we have an origin-destination (OD) matrix with elements 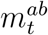 that represent the number of trips from district *a* to district *b* on day *t*. If a person makes a trip from district *a* to district *b* and comes back to district *a* within the same day, this is counted as two separate trips, one from district *a* to district *b*, and one from district *b* to district *a*. Let 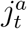; be the reported prevalence (active cases) of COVID-19 in district *a* on day *t*. We define the Pressure Score for district *b* from the past *n* days (non-inclusive of the current day) as:

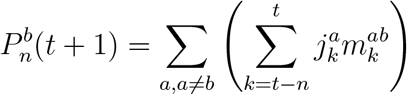

We normalize the Pressure Score for each district by that district’s population size so the score is calculated per 100,000 people. For our forecasts, we calculate the Pressure Score with *n* = 7 to reflect a week of travel, and often close to the incubation period of COVID-19 before it is observed and tested.

##### Internal Movement Score

The Internal Movement Score within a district reflects potential cases spreading within a district over the past *n* days. The diagonal of the OD matrix, given by 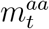, reflects the number of travelers who made a trip within district *a* on a certain day. We multiply this diagonal by daily COVID-19 prevalence within district *a* for the past *n* days (non-inclusive of the current day) and sum the results:

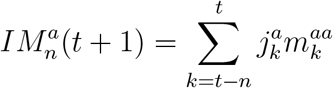

##### Excess Pressure and Internal Movement Scores

The Pressure and Internal Movement scores reflect absolute amounts of travel. However, we also want to understand how more or less travel compared to the norm affects the number of COVID-19 cases. We assume that the mobility data from February 2020 represents normal, pre-COVID-19 movement.

To compute the Excess Pressure score, we first compute excess daily OD matrices by subtracting from each day’s daily OD matrix the average OD matrix from that day of the week from February 2020. For example, if day *t* is a Friday, then we subtract the average OD matrix of the four Fridays in February 2020 (February 7, 14, 21, and 28) from 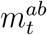. Let 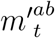 represent the excess number of trips compared to February 2020 from district *a* to district *b* on day *t*. If 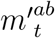 is positive, then there were more trips made on day *t* from district *a* to district *b* than the average for that day of the week in February 2020; and similarly, if 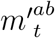 is negative, then there were fewer trips. The Excess Pressure and Internal Movement Scores are given by

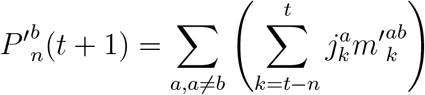

and

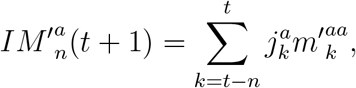

respectively.

##### Weekly Differenced Scores

To capture the weekly dynamics of mobility, we calculate weekly differenced values of the OD matrix by subtracting from each element of the OD matrix the corresponding value from seven days earlier. Let 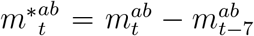 represent the weekly differenced number of trips from district *a* to district *b* on day *t*. If 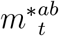 is positive, then there were more trips made on day *t* from district *a* to district *b* than there were on day *t* – 7; and similarly, if 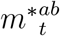 is negative, then there were fewer trips. The Weekly Differenced Pressure Score and Internal Movement Score are given by

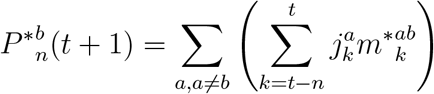

and

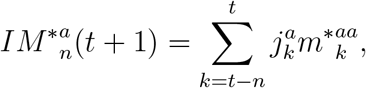

respectively.

#### 2.2.2 Model Outputs

For each district, we predict new cases per 100,000 people and the proportion of new tests that are positive, each averaged over the next seven days.

### 2.3 Predictive Modeling

An overview of our predictive modeling process is shown in Figure 2.

**Figure 2:**
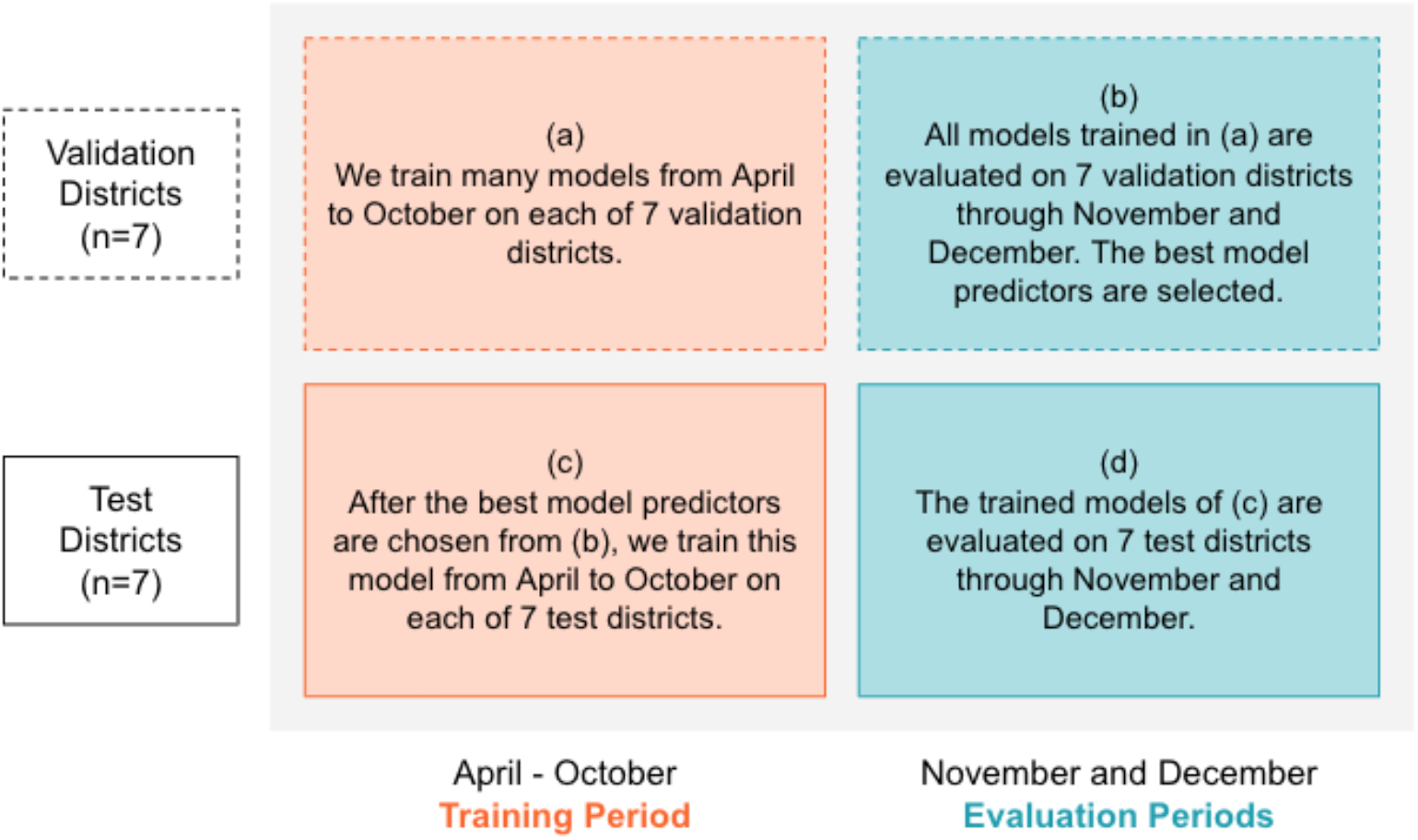
Overview of our predictive modeling process.

#### 2.3.1 Model type

Our predictive models use one-step-ahead linear regression. The model is retrained using daily updated data to make use of the most recent data in the predictions. Once all data is available to predict a given data point, we add that data point to our training set. Our problem is well suited to such a model because we have daily updated data for forecasting. We also considered deep learning models (details in Supplement C). We found that one-step-ahead linear regression generated the most accurate predictions (Table S.2).

Our linear regression model weights data points with a weekly exponential decay multiplier of 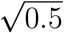 (half-life of two weeks). With this decay factor, the most recent week’s data has no decay, data from three weeks prior is weighted at half the most recent week, and from five weeks prior is weighted as one-fourth of the most recent week. We assign greater significance to more recent data points because the epidemic spread changes over time, as do social distancing regulations. We used the scikit-learn Python implementation (33) for the linear regression model.

#### 2.3.2 Time scales for prediction

Because our prediction problem involves time series data, we are restricted to training on previous data and evaluating our model on future forecasts. We define a training interval from April 6 – October 24, 2020, and two evaluation intervals from November 1 – 30, 2020, and from December 1 – 31, 2020 (Figure 2, horizontal axis). We began training on the earliest date for which each district had data, the earliest of which was April 6, the date when the first three districts in our dataset reported nonzero accumulated cases, tests, and recoveries. October 24 reflects an arbitrary “point of adoption” for our models, after which the model has learned enough from the training interval to forecast into the future. To prevent data leakage, we ensure a difference of seven days between the training interval and evaluation interval to account for the fact that our prediction encodes information for the following seven days. The November and December evaluation intervals are separate because the November evaluation interval includes realized mobility data, whereas the December evaluation interval uses perturbed estimates of mobility data.

#### 2.3.3 Model scope

We randomly sorted the 14 districts into 7 validation districts and 7 test districts (Figure 2, vertical axis). Models for the validation districts were trained on the training interval, then evaluated on the evaluation interval to determine the best model attributes. We evaluate the model and report results on the test districts over the evaluation interval. We used the same model predictors for all districts, chosen by best performance on the validation districts. Each district has its own model to be trained and evaluated on, so the weights of each district’s model are tuned according to the specific dynamics in that district.

#### 2.3.4 Model evaluation

All models were evaluated by mean squared error (MSE), averaged over all districts in each evaluation set. MSE measures the average of the squares of the errors, or the difference between the actual (*Y*) and predicted (*Ŷ*) values of *n* data points, as follows:

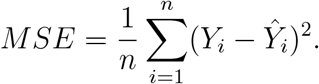

We report root mean squared error (RMSE, the square root of the MSE) because it is interpretable in terms of measurement units of a certain number of cases per day (for new cases) or a percentage (test positivity rate). Validation RMSE reflects performance on the 7 validation districts through November and December (Figure 2b), and test RMSE reflects performance on the 7 test districts through November and December (Figure 2d).

### 2.4 Severity Categorization

We use a decision rule to classify the predicted severity of a COVID-19 outbreak in each district over the following week. We report results for each district-date pair. The scheme categorizes new cases and the test positivity rate into five tiers (Table 1). From the validation districts over the training interval (Figure 2a), we computed the 20th, 40th, 60th, and 80th quantiles of new cases and the test positivity rate, respectively. These values define the tiers, which represent quintiles within our training set. We note that these tiers were developed with some hindsight bias, assuming that the large spike Israel faced in the late summer of 2020 is representative of future spikes they could face.

**Table 1:** Severity categorization scheme defined by the quintiles of the training set.

| Severity | New Cases | Test Positivity Rate |
| --- | --- | --- |
| Tier 5 | $\geq 33.0$ | $\geq 10.0\%$ |
| Tier 4 | 16.0 – 32.9 | 7.0 – 9.9% |
| Tier 3 | 9.0 – 15.9 | 5.0 – 6.9% |
| Tier 2 | 3.0 – 8.9 | 2.5 – 4.9% |
| Tier 1 | $< 3.0$ | $< 2.5\%$ |

We compare results using our categorization scheme to results using a four-tier system used by the State of California, which categorizes COVID-19 spread as “minimal,” “moderate,” “substantial,” or “widespread” based on new cases and the test positivity rate (Table S.1) (34). This is similar to the four-tier system used in Israel, which calculates a score based on new cases, the test positivity rate, and the daily growth rate of active cases (35).

We use magnitude accuracy (MA) to evaluate the performance of our predictions when classified into severity tiers. All magnitude accuracy metrics reflect performance on the 7 test districts through November and December (Figure 2d). We measure the proportion of days for which the predicted and actual values fall within the same tier and for which the predicted value falls within one tier of the actual value’s tier. We also report confusion matrices that visualize the accuracy of our predictions as classified into the tiers.

### 2.5 Selection of model predictors

To inform the selection of predictors for our models, we explored correlations within the health data and correlations between the health data and the non-COVID-related data such as mobility and socioeconomic data (details in Supplement B). We calculated linear correlation scores (Pearson’s *r* score) over two periods: the period for which we have actual mobility data (April 6 – November 30, “actual mobility period”) as well as the period of the entire dataset (April 6 – December 31, “extended mobility period”). The extended mobility period includes the month of December and uses perturbed estimates of mobility during that month. Figures S.2 and S.3 show key correlations for new cases and the test positivity rate, respectively, over the actual mobility period and the extended mobility period. The correlation coefficients over these two evaluation sets are comparable, indicating that our perturbed mobility estimates are reasonable.

We defined 54 unique combinations of predictors from the predictors defined in Section 2.2.1. We considered the best set of predictors that did not include mobility data, evaluated based on RMSE, as the baseline. Then we assessed whether including mobility predictors improved performance in terms of both RMSE and MA. We ranked the models that included a mobility predictor and had lower RMSE than the baseline in increasing order of RMSE. Starting from the model with the lowest RMSE, if the subsequent model had higher MA, we selected that model instead. We stopped iterating when both RMSE increased and MA decreased and report this as the best model that includes a mobility predictor. We selected features in this way rather than performing ANOVA or selecting features through Lasso regression because this selection method is transferrable to other model types (Supplement C).

### 2.6 Ethics Approval

The IRB chair of Tel Aviv University, Prof. Meir Lahav, determined on March 24, 2020 that an IRB approval is not needed for this study. We received consent from the data provider to use the aggregated and anonymized human mobility data in the way it is used in this study.

## 3 Results

### 3.1 Characteristics of Districts

Table 2 presents descriptive statistics for the 14 included districts, stratified by whether the districts were randomly sorted into the validation or test sets. On average, the validation districts were less populated than the test districts, and their population was younger and more impoverished. The validation districts had a greater number of daily new cases, but fewer new tests, so the proportion of COVID-19 tests that were positive was slightly higher in the validation districts than in the test districts. The validation districts had more travel from outside the district but less travel within the district than the test districts. The overall median age of 29.7 years in our dataset is close to the reported median Israeli age of 30.5 years (36).

**Table 2:** Descriptive characteristics of the districts in our dataset. Population size, socioeconomic score, and median age are fixed over time. Active cases, new cases, new tests, test positivity rate, pressure, and internal movement reflect daily values per 100,000 people. Pressure score is calculated daily.

|  | All Districts |  | Validation Districts |  | Test Districts |  |
| --- | --- | --- | --- | --- | --- | --- |
| # Districts | 14 |  | 7 |  | 7 |  |
| Median earliest day in dataset | April 23 |  | April 23 |  | April 20 |  |
| Characteristic | Mean | 95% CI | Mean | 95% CI | Mean | 95% CI |
| Population | 590,624 | 386,058-795,190 | 445,902 | 126,851-764,952 | 735,346 | 436,955-1,033,738 |
| Socioeconomic Score | 3.96 | 3.03-4.88 | 3.44 | 1.89-4.99 | 4.48 | 3.13-5.82 |
| Median Age | 29.7 | 27.7-31.7 | 28.2 | 25.4-31.0 | 31.2 | 28.1-34.2 |
| Active Cases | 266.4 | 257.5-275.3 | 303.1 | 287.6-318.6 | 229.5 | 220.9-238.2 |
| New Cases | 21.78 | 20.87-22.69 | 24.96 | 23.38-26.54 | 18.59 | 17.71-19.47 |
| New Tests | 366.6 | 354.4-378.9 | 362.5 | 343.1-381.9 | 370.8 | 355.8-385.8 |
| Test Positivity Rate | 0.0516 | 0.0502-0.0530 | 0.0564 | 0.0542-0.0586 | 0.0467 | 0.0451-0.0484 |
| Pressure Score | 79.1 | 76.8-81.4 | 102.2 | 98.2-106.2 | 55.9 | 54.1-57.7 |
| Internal Movement Score | 210.3 | 202.0-218.5 | 178.9 | 171.3-186.4 | 241.8 | 227.3-256.3 |

### 3.2 Predictive Modeling

Because the state of the epidemic differed between November and December, we selected the best predictors over both evaluation periods. In November, there was a lull in cases in most districts. Then, in most districts, there was a spike in cases near the end of December. An ideal model would be able to predict both scenarios (low cases and a spike in cases) with equal ability, rather than specializing for one task. Therefore, we weighted the evaluation periods equally when selecting predictors. Figure S.4 shows the tradeoff between November and December in the performance of different sets of predictors in predicting new cases and the test positivity rate.

Models using mobility data outperformed models that did not use mobility data, reducing RMSE by 17.3% when predicting new cases and by 10.2% when predicting the test positivity rate (Table 3).

**Table 3:** Performance of one-step-ahead linear regression in predicting new cases and test positivity rate, averaged over the next 7 days, reported as RMSE (root mean squared error).

| Prediction Task | Best Predictors | RMSE |  |
| --- | --- | --- | --- |
|  |  | *Validation Districts* | *Test Districts* |
| New Cases | With mobility | 6.825 | 4.812 |
|  | Without mobility | 8.205 | 5.817 |
| Test Positivity Rate | With mobility | 1.02% | 0.79% |
|  | Without mobility | 1.21% | 0.88% |

#### 3.2.1 RMSE for Predicting New Cases

The best set of predictors for new cases comprised 1-day lag of past 7-day average incidence and the Excess Internal Movement Score. This model had an RMSE of 6.825 new cases per day on the validation set and 4.812 new cases per day on the test set (Table 3). Figure 3 shows predictions of new cases for the validation and test districts. Despite significant fluctuations in the true number of new cases in the following week, our model accurately predicts levels and trends in new cases a week in advance.

**Figure 3:**
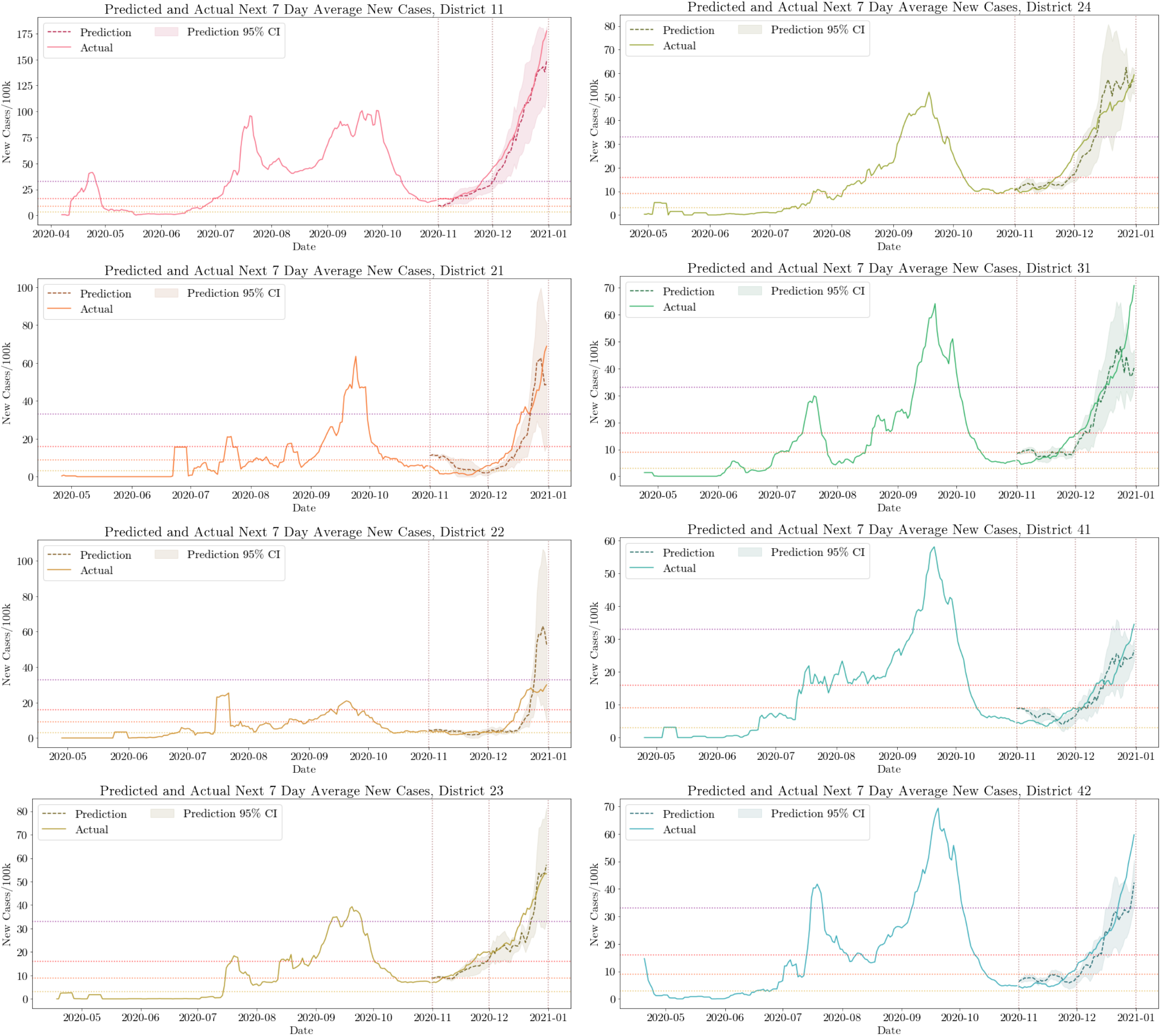
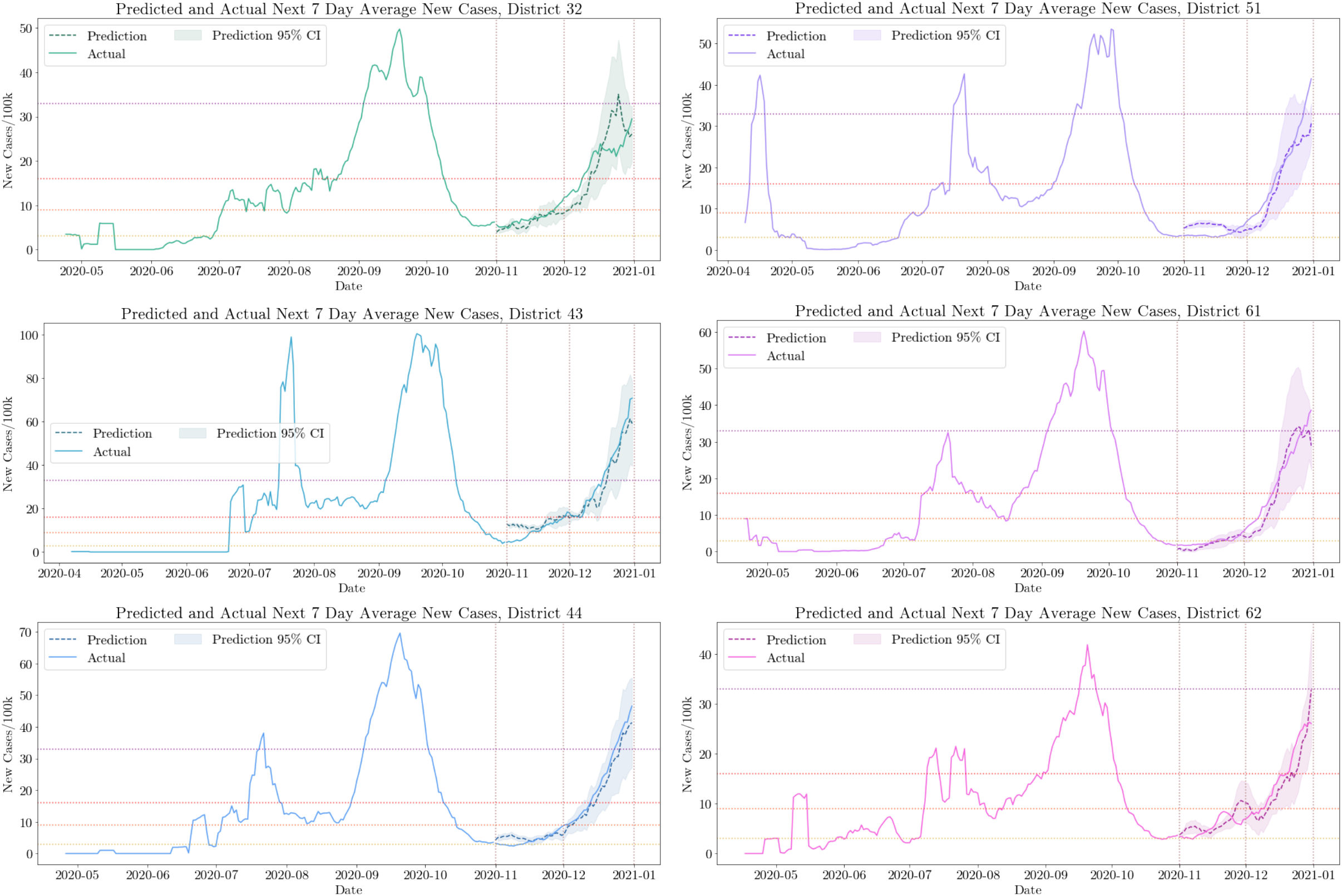
Predicted and actual next 7-day average new cases in the seven validation districts (left) and the seven test districts (right). Our predictions, made a week in advance, are shown as a dashed, darker line. The 95% confidence interval, computed using the past 14-day average standard deviation in predicted new cases, is shown in gray shading. Vertical dotted lines delineate the separate prediction intervals of November and December. The lower bound of Tiers 2-5 used for classifying the level of epidemic severity from Table 1 are shown as horizontal dotted lines.

#### 3.2.2 RMSE for Predicting the Test Positivity Rate

The best set of predictors for the test positivity rate comprised 3-days lag of past 7-day average test positivity rate and the Excess Internal Movement Score. This model had an RMSE of 1.02% and 0.79% on the validation and testing districts, respectively.

### 3.3 Severity Categorization

Magnitude accuracy for the prediction of new cases and the test positivity rate is shown in Table 4. Each of the 7 testing set districts contributes 61 data points (30 from November and 31 from December), yielding a sample size of 427. The use of mobility data improved magnitude accuracy in predicting new cases by 24% and magnitude accuracy in predicting the test positivity rate by 5%. Test positivity rate predictions are less affected by mobility because the variance in the daily number of new tests affects the results more.

**Table 4:** Performance of our best performing predictions of new cases and the test positivity rate as classified into tiers by our categorization scheme, reported as magnitude accuracy (MA).

| Prediction Task | Best Predictors | Magnitude Accuracy |  |
| --- | --- | --- | --- |
|  |  | *Validation Districts* | *Test Districts* |
| New Cases | With Mobility | 0.637 | 0.775 |
|  | Without Mobility | 0.564 | 0.623 |
| Test Positivity Rate | With Mobility | 0.630 | 0.820 |
|  | Without Mobility | 0.588 | 0.778 |

#### 3.3.1 Magnitude Accuracy for New Cases

Confusion matrices are shown in Figure 4. Our severity categorization scheme for new cases never misidentifies a minor outbreak (Tiers 1-3) as a very severe one (Tier 5) or a very severe outbreak (Tier 5) as a minor one (Tiers 1-3). These are respectively visualized through zeroes in the rightmost column, top three squares, and zeroes in the bottom row, leftmost three squares of Figure 4a.

**Figure 4:**
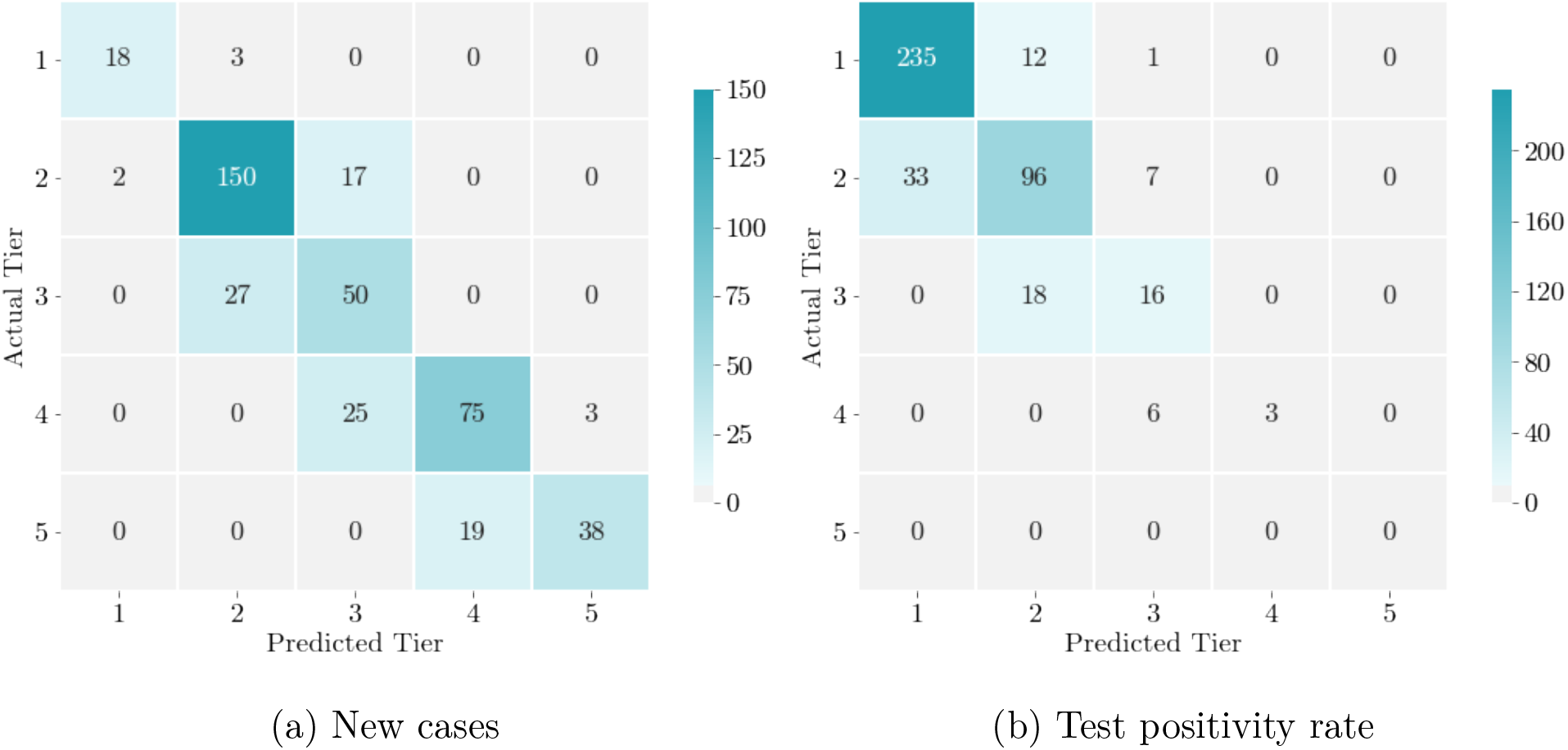
Confusion matrices illustrating the accuracy of the severity categorization of our model’s predictions of new cases and proportion of COVID-19 tests that are positive from November 1 to December 31, 2020 (*n* = 427). Columns represent the model-predicted severity for the following week, categorized into the five tiers, where Tier 1 is the least severe and Tier 5 is the most severe, and rows represent actual values of epidemic severity for the following week.

We accurately predicted 38 of 57 district-date instances where next week truly fell into Tier 5, and 38 of the 41 district-dates that were predicted to be in Tier 5 were truly in Tier 5, leading this tiering system to have a sensitivity of 0.67 and a positive predictive value of 0.93 for new cases being in the most severe tier.

The overall proportion of days for which the predicted and actual average new cases fall in the same severity tier is 0.775, which is significantly higher than would be achieved by random guessing (.20). The tier-specific classification accuracies are 0.86, 0.89, 0.65, 0.73, 0.67, for Tiers 1-5 (Figure 4a). Misclassified district-date pairs always fell within one tier of the actual severity tier; thus, the accuracy of our model in predicting severity within one tier is 1. This is reflected in the dark color of the diagonal shifted by one square in each direction in Figure 4a.

#### 3.3.2 Magnitude Accuracy for the Test Positivity Rate

Magnitude accuracy using the test positivity rate tiers is 0.820 when using mobility data (Table 4). The tier-specific classification accuracies vary from 0.95 for Tier 1 to 0.71, 0.47, 0.33 for Tiers 2-4, respectively (Figure 4b).

Our tiers were selected based on test positivity rates that occurred between April to October. In November and December, most districts had lower daily test positivity rates. The higher test positivity rate at the beginning of the pandemic is indicative of the limited test supply at that time. No district-date pairs truly fell into Tier 5 during our testing period.

During this period, 58% of district-date pairs in our test districts fell into Tier 1, pictured by the dark blue squares at the top left corner of Figure 4b. Because of this skew in the data, the predicted tier understates the actual tier more often than not. Excluding districts that truly fell into Tier 1 over the next week, 31.8% of districts were classified as one severity tier below their true tier, which is almost half as many as were classified into their true tier (64.2%, Figure 4b). Nonetheless, our model has high magnitude accuracy in predicting the test positivity rate in each district.

#### 3.3.3 Magnitude Accuracy when Using California Severity Categorization

Figure S.5 presents confusion matrices for our predictions when using California’s categorization scheme for epidemic severity. Magnitude accuracy for our categorization scheme is 0.775 for predicting new cases and 0.820 for predicting the test positivity rate. When using the less balanced California tiers, magnitude accuracy for these two quantities decreases to 0.648 and 0.765, respectively. Additionally, an outbreak is categorized as “widespread” more than half of the time when using the California tiers, whereas our categorization allows for a more detailed distinction between levels of outbreak spread.

## 4 Discussion

In today’s interconnected world, human mobility data in the form of cell phone data can provide valuable insight into human behavior. We have shown that aggregate and anonymized cell phone mobility data can be used to improve the prediction of COVID-19 outbreaks, measured as daily new cases and the test positivity rate. Our best model’s predictors consisted of 1-day lag of past 7-day average new cases or 3-days lag of the test positivity rate, respectively, along with the Excess Internal Movement Score. Our forecasting approach allows for early identification of regions that may develop outbreaks in the coming week, potentially allowing policy measures to be targeted to the regions where they would be the most effective. Additionally, we found that a balanced tiering system for categorizing outbreak severity (based on fractiles of previously observed data) allows for more accurate prediction than an unbalanced tiering system.

Our analysis has several limitations. We assumed that both the mobility and the health data were relatively accurate estimates of the true amounts of travel and prevalence of COVID-19 in a region, respectively. If the health data for a given district is skewed due to selection bias in who receives tests, forecasts for other districts would be affected through the mobility data. Districts were included in our dataset only when one statistical region within the district reported at least 15 accumulated cases, tests, and recoveries. Each time a statistical region started to be documented in the health dataset, our dataset experienced an increase in the number of cases that may not reflect an actual outbreak. Future work could develop methods to impute these missing values with constraints based on the total number of reported cases on a day. Smoother data would aid predictions of actual outbreaks as models would be less likely to overfit to random noise in the dataset. Our analysis predicts new cases based on information about known cases and does not take into account cases that were never detected (e.g., asymptomatic cases). Future work could develop methods for adjusting predictions to accurately account for undetected cases.

Our models predict new cases more accurately than the test positivity rate. This is because the daily changing sample sizes make it hard to consider the test positivity rate as a consistent stochastic process or to draw conclusions based on the test positivity rate’s patterns. For example, an increase in the daily test positivity rate does not necessarily indicate a worsening situation in a region if the sample size on this date is relatively small, and vice versa. Further research could investigate the use of predictors that take into account sample size, if known.

Cell phone mobility data of the type we used may not be available in other settings. Future work could use Google Community Mobility data, available for most countries online (37). This dataset provides information about mobility trends across different categories of places, including retail, grocery, parks, transit, workplaces, and residential areas, measured as changes from a baseline in January 2020. The Community Mobility data is similar to our Excess Pressure and Internal Movement scores, both of which we found useful in predicting new cases. Although not as granular as the data we used (for example, the Community Mobility data divides Israel into six regions), such mobility data could still be useful in predicting COVID-19 spread.

Our work shows that anonymized, aggregated human mobility data can improve the prediction of when and where COVID-19 outbreaks are likely to occur. We merged anonymized country-level data from different sources (e.g., health data, socioeconomic data, cell phone mobility data) to develop a simple and accurate method for predicting weekly new COVID-19 cases and the test positivity rate. Accurate prediction of outbreak severity allows for better allocation of resources (e.g. vaccines, medical staff, targeting of lockdown policies) at a district level. Our method provides a useful tool for government decision makers, particularly in the post-vaccination era, when focused interventions are needed to contain COVID-19 outbreaks while mitigating the collateral damage of more global restrictions. The methods we have developed can also be applied to predict outbreaks of other communicable diseases such as influenza, measles, and SARS.

## Data Availability

The datasets analysed during the current study are available from the corresponding author on reasonable request. Pending peer review, all data and code will be made publicly available online.

## Acknowledgements

This work was supported by a Koret Foundation gift for Smart Cities and Digital Living.

## Declarations

### Authors’ contributions

Conceptualization: GG, DY, MLB

Methodology: GG, IBG, DY, MLB

Investigation: GG, YD, MY

Visualization: GG, MY, DY, MLB

Supervision: IBG, DY, MLB

Writing—original draft: GG, MLB

Writing—review & editing: GG, YD, MY, IBG, DY, MLB

