## Supplement for "Early Detection of COVID-19 Outbreaks Using Human Mobility Data"

#### A Synthetic Mobility Data for December 2020

Because the mobility dataset ended on November 30, 2020, we generated synthetic mobility data for December 1-31, 2020. For each date in December, we identified dates that had similar mobility restrictions earlier in the pandemic ("similar dates"). We validated the similarity of restrictions between dates by a routine index presented by one of the leading news companies in Israel (38). The index uses the number of commuters on public transportation, the unemployment rate, and the rate of expenses, among other factors, to represent the similarity between mobility on any given day and a pre-COVID-19 "routine" day, where a score of 100% reflects routine mobility before the pandemic. The first week of December was similar to the last week of November, the middle three weeks of December were similar to July, and the last five days of December were similar to the third and fourth weeks of September. We matched each date in December to the corresponding similar date(s) by the day of the week. Then, we estimated mobility data for the December dates by perturbing averaged mobility data over their similar dates. We either added or subtracted a random noise element centered around 0 with a standard deviation of 25% of the similar dates' values, based on whether the restrictions were exactly the same (positive or negative perturbation) or less severe (positive perturbation; more mobility). For example, if all restrictions were the same on days A and B, except that schools were open on day B, we would apply a positive perturbation to day A's mobility data to estimate mobility on day B.

#### B Correlations between Model Inputs and Outputs

Lagged health features were most strongly correlated with the same health features in the next 7 days. These correlations are independent of the mobility data. A past week's 7-day average of new cases was strongly correlated with the following week's 7-day average of new cases ($r = 0.86$ and $r = 0.87$ during the actual and extended mobility periods, respectively; Figure S.2), and a past week's test positivity rate was strongly correlated with the next week's average test positivity rate ($r = 0.65$ and $r = 0.77$ during the actual and extended mobility periods, respectively; Figure S.3). Because of the strong correlations between lagged health features and our intended predictors, we included 1-day, 3-day, and 6-day lagged health features as potential predictors for each prediction task.

We found lagged correlation between inter-district travel and COVID-19 incidence. In particular, if district A has a high number of COVID-19 cases and there has been significant travel from district A to district B over the past week, then we find that district B will have a relatively high number of new COVID-19 cases in the following week. The Pressure Score for a district from the past 7 days was correlated with new cases in the district averaged over the next 7 days, with $r = 0.47$ (actual mobility period) and $r = 0.66$ (extended mobility period). The Internal Movement Score for a district from the past 7 days was also correlated with new cases averaged over the next week, with $r = 0.53$ (actual mobility period) and $r = 0.36$ (extended mobility period). Similarly, the average test positivity rate over the next week was correlated with the Pressure Score from the past 7 days, with $r = 0.26$ (actual mobility period) and $r = 0.34$ (extended mobility period), and also correlated with the Internal Movement Score from the past 7 days with $r = 0.45$ (actual mobility period) and $r = 0.24$ (extended mobility period). We therefore included the Pressure and Internal Movement Scores, alone and combined, as potential predictors for all prediction tasks.

Negative correlations between a week's new COVID-19 cases and the previous week's differenced and Excess Pressure and Internal Movement Scores reflect the effects of lockdowns and other policy measures implemented to reduce COVID-19 spread. A greater reduction in mobility compared to the February 2020 baseline, as reflected by the Excess Internal Movement and Excess Pressure Scores, was correlated with a reduction in new cases over the next 7 days ($r = -0.30$ for both Excess Pressure and Internal Movement, over the actual mobility period, and $r = -0.35$ and $-0.32$, respectively, over the extended mobility period). The Excess Internal Movement and Excess Pressure Scores were also correlated with a reduction in test positivity rate over the next 7 days over both evaluation periods ($r$ scores of $-0.40$, $-0.50$, $-0.30$, $-0.31$, respectively). Differenced Pressure and Internal Movement Scores produced similar results of slightly smaller magnitude. As we did for the Pressure and Internal Movement Scores, we considered the excess and differenced mobility predictors separately and together as predictors for all prediction tasks.

Because we predicted cases within each district separately, we did not include socioeconomic score or median age, which are constant factors, as potential features when predicting new cases and the test positivity rate. However, we note that socioeconomic score was negatively correlated with the test positivity rate: poorer districts tended to have higher test positivity rates ($r = -0.39$ and $r = -0.23$ during the actual and extended mobility periods, respectively). Median age was correlated with new cases ($r = -0.47$ and $r = -0.20$ during the two periods, respectively) but socioeconomic score was not correlated with new cases ($r = -0.07$ and $r = -0.03$ during the respective periods).

#### C Predictions: Other Model Types

In addition to one-step-ahead linear regression models, we tested deep learning models in the form of recurrent neural networks (RNNs). Though autoregressive moving average (ARIMA) models can accurately forecast COVID-19 in other situations (11–13), the high variance and lack of weekly repeating patterns in daily new cases (Figure S.1) and tests made ARIMA models impractical for our dataset.

RNNs capture temporal behavior in data and can use past steps as input to generate output sequences (39). One RNN architecture that has had success in clinical time series applications, such as accurately forecasting outpatient clinic demand (40, 41), is the long short-term memory (LSTM) architecture. Past literature has explored using RNNs and LSTMs for predicting accumulated cases of COVID-19 (13–17). Our network structure composed an RNN with one hidden LSTM layer of 8 units; this structure performed the best on the validation districts. We trained our LSTM for 1,000 epochs with an early stopping function to pause training if validation mean squared error had not decreased for 5 epochs. When training the LSTM, we took out a validation period from our training interval from September 16 – October 24, 2020. We implemented this model with TensorFlow (42).

We evaluated these models' predictions of new cases during the period for which we had actual mobility data, November. One-step-ahead linear regression models outperformed the RNN (Table S.2), most likely because RNNs require larger amounts of data to be trained optimally.

### S Supplemental Figures and Tables

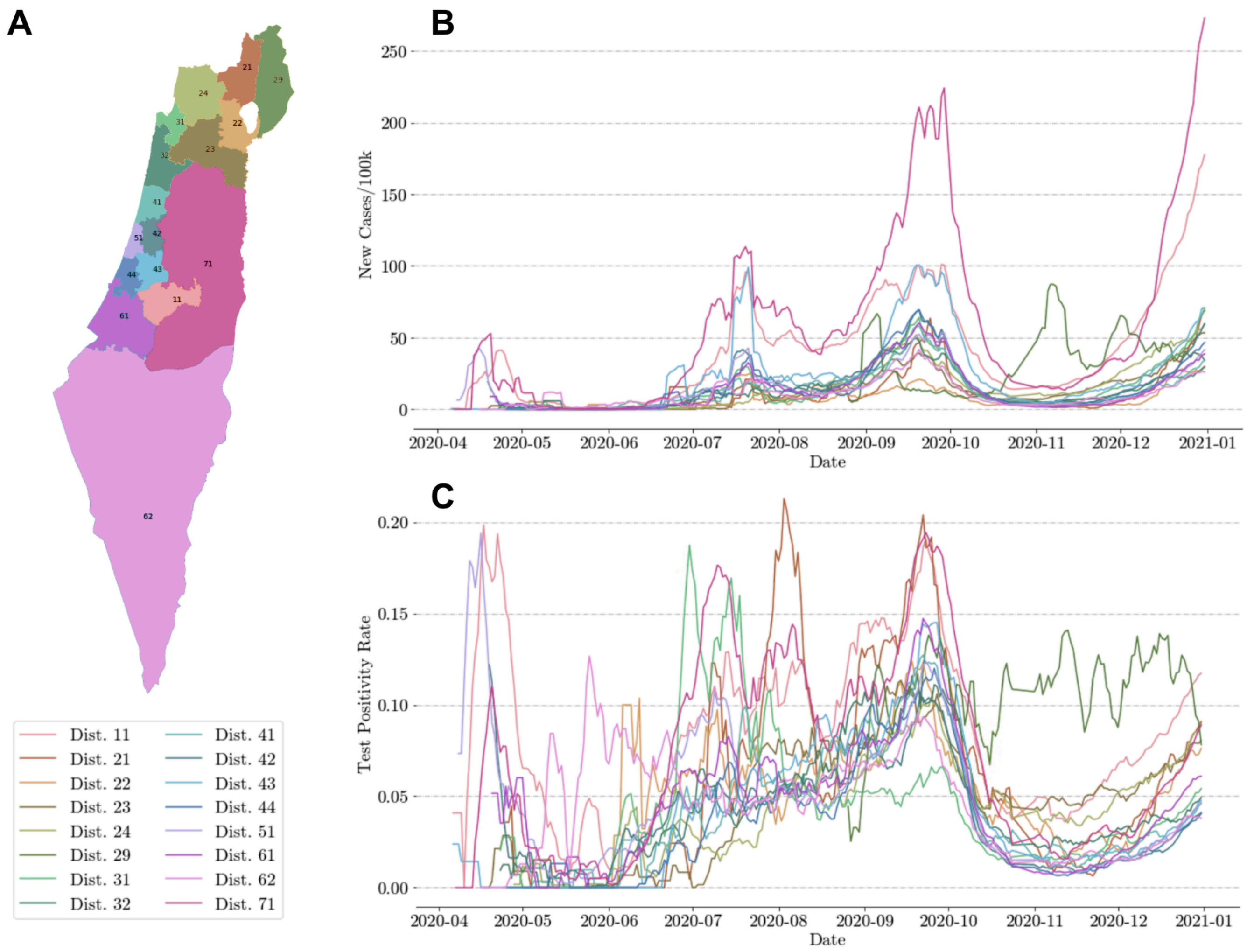


Figure S.1: (A) A map showing each of the 16 districts in Israel (B) Seven day rolling average of new cases per 100,000 people for each of the districts (C) Seven day rolling average of test positivity rate for each of the districts.

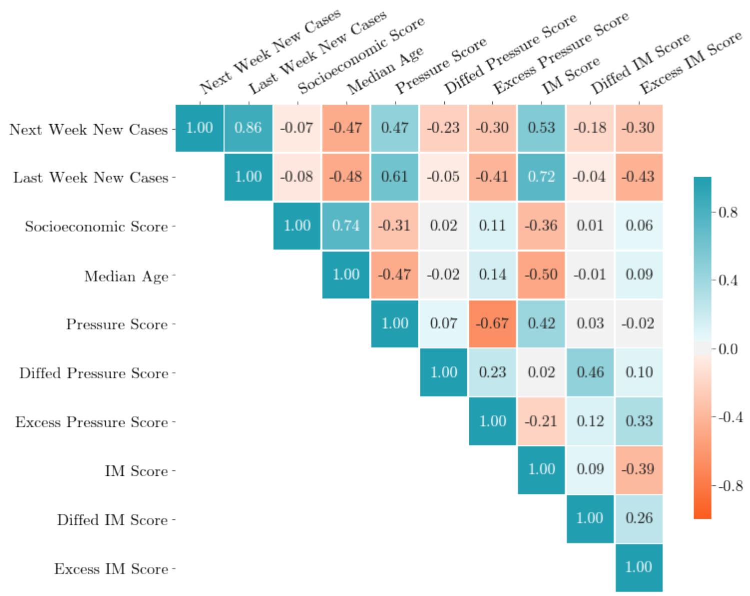


(a) Pearson's $r$ correlation scores between new cases and socioeconomic and mobility data during the actual mobility period.

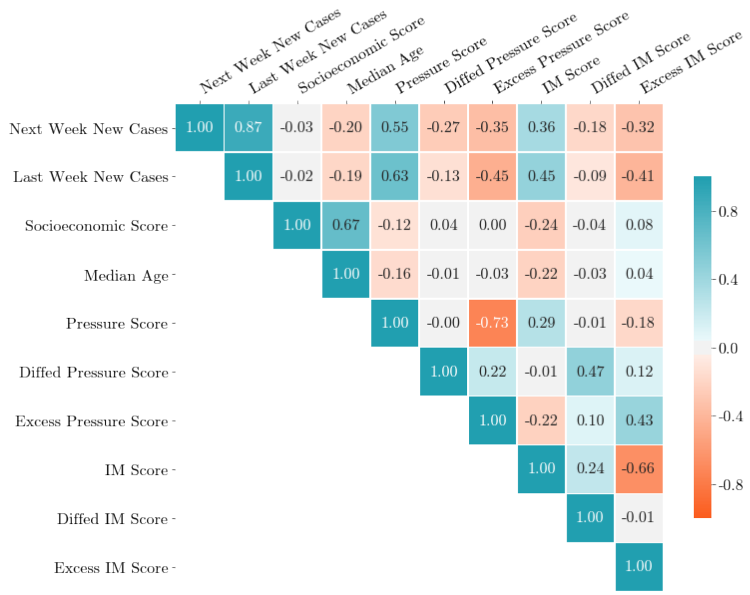


(b) Pearson's $r$ correlation scores between new cases and socioeconomic and mobility data during the extended mobility period.

Figure S.2: Heatmaps showing correlation coefficients between new cases averaged over the next seven days and health, demographic, and mobility characteristics for the 14 districts for the actual and extended mobility periods. New cases and the Pressure and Internal Movement Scores are per 100,000 people.

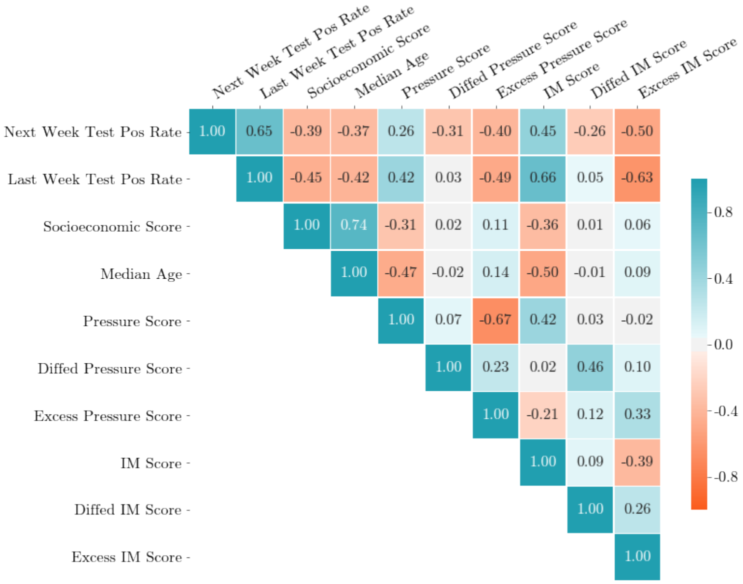


(a) Pearson's $r$ correlation scores between test positivity rate and socioeconomic and mobility data during the actual mobility period.

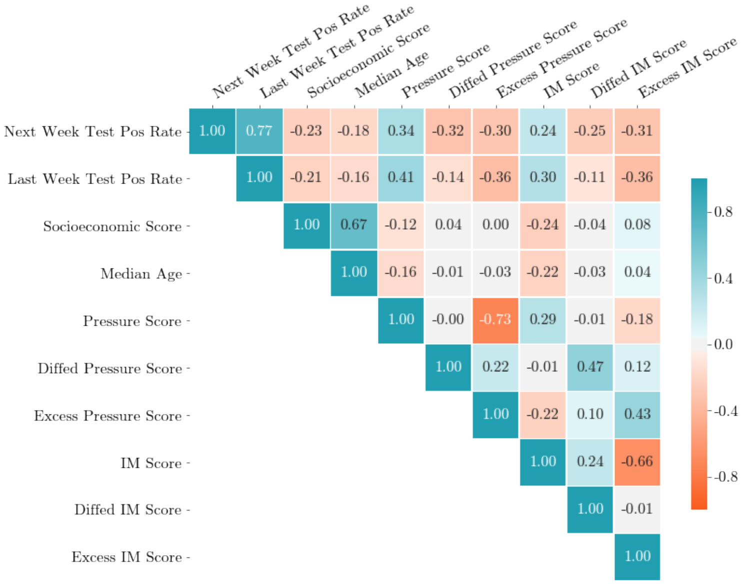


(b) Pearson's $r$ correlation scores between test positivity rate and socioeconomic and mobility data during the extended mobility period.

Figure S.3: Heatmaps showing correlation coefficients between average test positivity rate in the next seven days and health, demographic, and mobility characteristics for the fourteen districts over the actual and extended mobility periods. Tests and the Pressure and Internal Movement Scores are per 100,000 people.

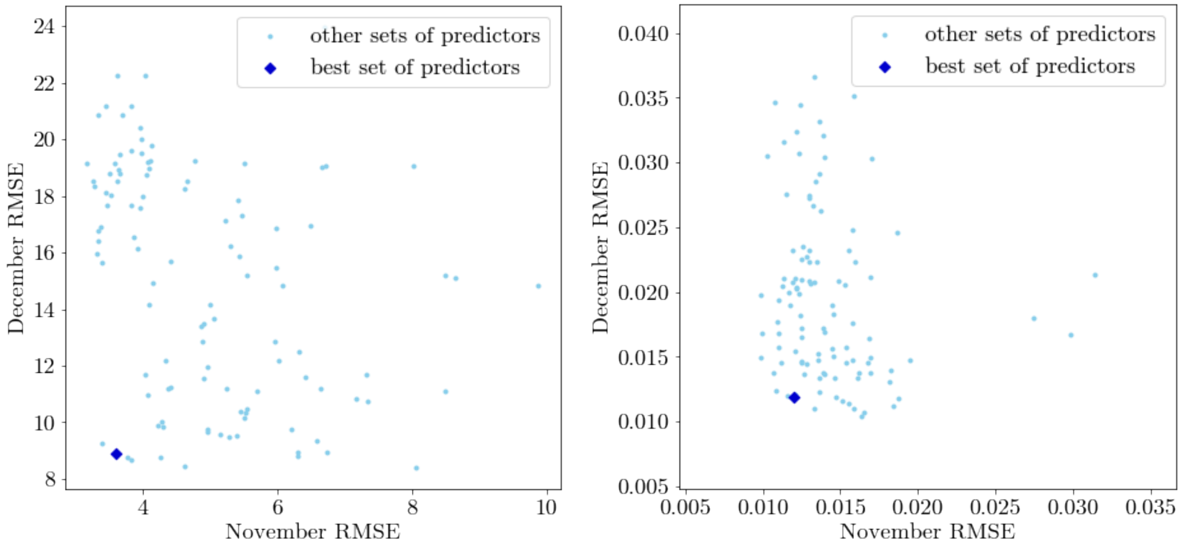


(a) New cases (b) Test positivity rate

Figure S.4: Performance of different sets of predictors in predicting (a) new cases and (b) test positivity rate measured as root mean squared error (RMSE) for November (x-axis) and December (y-axis). The set of predictors that minimized average RMSE over each month is shown as a dark blue diamond and is near the bottom left corner of each plot.

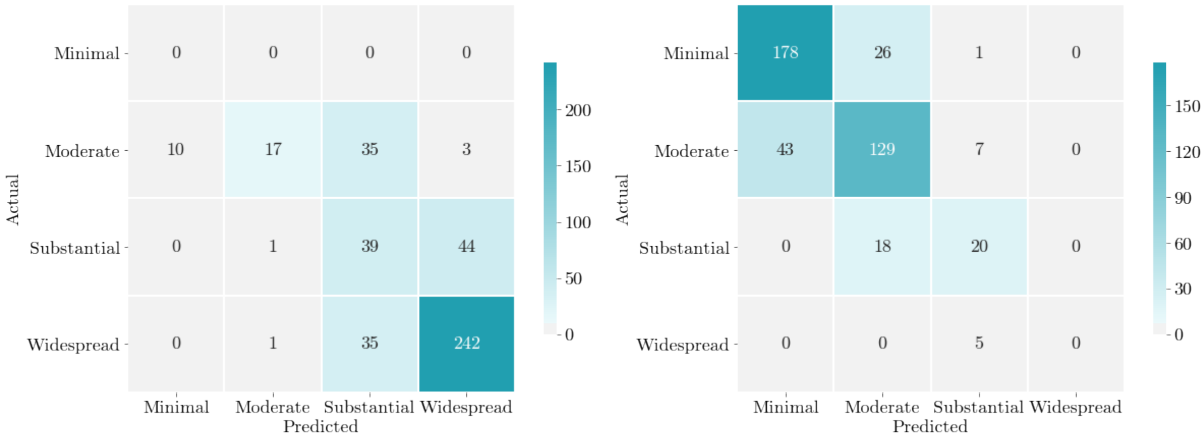


(a) New cases, CA categorization (b) Test positivity rate, CA categorization

Figure S.5: Confusion matrices for the best prediction model over the 7 testing districts ($n = 427$) for the California (CA) severity categorization scheme. Tiers are defined by (a) new cases and (b) test positivity rate that are positive from November 1 to December 31, 2020.

| **Magnitude** | **New Cases** | **Test Positivity Rate** |
| --- | --- | --- |
| Widespread | $\geq 7.0$ | $\geq 8.0\%$ |
| Substantial | 4.0 – 6.9 | 5.0 – 7.9% |
| Moderate | 1.0 – 3.9 | 2.0 – 4.9% |
| Minimal | $< 1.0$ | $< 2.0\%$ |

Table S.1: Tiers used by the State of California to classify COVID-19 outbreak magnitude.

| **Model Architecture** | **RMSE** |
| --- | --- |
| Linear Regression: One-step-ahead | 3.617 |
| RNN | 5.957 |

Table S.2: Comparison of the models tested when predicting new cases, by root mean squared error (RMSE) on validation districts over the evaluation period of November only.
